# Per- and Poly-fluoroalkyl Substances (PFAS) in Circulation in a Canadian Population: Their Association with Serum Liver Enzyme Biomarkers and Piloting a Novel Method to Reduce Serum PFAS

**DOI:** 10.1101/2024.08.02.24311171

**Authors:** Jennifer J. Schlezinger, Anila Bello, Kelsey M. Mangano, Kushal Biswas, Paridhiben P. Patel, Emily H. Pennoyer, Thomas M.S. Wolever, Wendy J. Heiger-Bernays, Dhimiter Bello

**Author notes:** **Corresponding author:** Jennifer J. Schlezinger, Ph.D. Boston University School of Public Health Dept. of Environmental Health 715 Albany Street, R-405 Boston, MA 02118.

## Abstract

Extensive use of per- and polyfluoroalkyl substances (PFAS) has resulted in their ubiquitous presence in human blood. PFAS exposures have been associated with multiple adverse human health effects including increased risk of liver damage, elevated serum lipids, impaired vaccine response, adverse birth outcomes and cancer. Biomonitoring studies have focused on measuring long-chain PFAS, but these are being replaced by shorter chain PFAS and PFAS with alternative structures, resulting in incomplete understanding of human exposures. Here, we take advantage of serum samples collected as part of a clinical trial testing the efficacy of a dietary fiber intervention to reduce serum cholesterol to investigate exposure to legacy and replacement PFAS chemicals in Canadian participants. Serum samples were collected from 72 participants in 2019-2020 at baseline and after 4 weeks of the intervention and were analyzed for 17 PFAS species. The highest geometric mean concentrations of PFAS measured at baseline corresponded to PFOSA (7.1 ng/ml), PFOS (4.2, ng/ml), PFOA (1.8 ng/ml) and PFHxS (1.3 ng/ml). Short chain PFAS including PFBuA, PFHxS and/or PFHpA were detected in 100% of participants and GenX was detected in 70% of participants. Analyses of associations between PFAS serum concentrations and biomarkers of adverse health outcomes showed the PFBuA, PFHxA, PFDA and PFOSA were associated with higher serum gamma-glutamyl transferase concentrations but not with measures of serum total or low-density lipoprotein cholesterol. Comparison of PFAS concentrations at baseline and after a 4-week follow-up showed that total PFAS concentrations decreased in both the control and cholesterol intervention groups. However, the suite of PFAS of concern identified by the United States National Academies of Sciences, Engineering, and Medicine, significantly decreased only in the cholesterol intervention group. This observation suggests that a dietary fiber intervention may reduce PFAS body burden, but future intervention studies need to control for PFAS exposure sources and extend beyond 4 weeks. Overall, the results show that exposures to short-chain and alternative PFAS are common in this Canadian population.

## 1. Introduction

Per- and polyfluoroalkyl substances (PFAS) are a large class of mostly unregulated pervasive, mobile, and bioaccumulative chemicals, to which people are exposed daily from food, drinking water, consumer products and occupational activities (EFSA, 2018; Kim et al., 2019; Makey et al., 2017). Recent analyses show that environmental contamination by and human exposure to legacy PFAS remain a global problem. By “legacy” we mean longer-chain perfluoroalkyl substances and their precursors historically used for commercial and industrial uses, including aqueous film forming foam (AFFF), processing aids in manufacturing, and food contact materials (Glüge et al., 2020). Numerous legacy PFAS such as perfluoroheptanoic acid (PFHpA), perfluorooctanoic acid (PFOA), perfluorononanoic acid (PFNA), perfluorodecanoic acid (PFDA), perfluoroundecanoic acid (PFUdA), perfluorododecanoic acid (PFDoA), perfluorohexane sulfonic acid (PFHxS), and perfluorooctane sulfonic acid (PFOS) are commonly detected in drinking water, fresh water fish, food packaging, air and human sera (Cookson and Detwiler, 2022; Kurwadkar et al., 2022). It is estimated that 200 million people in the United States alone rely on tap water contaminated with PFAS (Andrews and Naidenko, 2020). Where analyzed, multiple PFAS are universally detected in the serum of human populations (De Silva et al., 2021). In addition to drinking water and food exposures, there is significant occupational exposure to PFAS; blood levels of PFAS are elevated in firefighters, ski wax technicians and PFAS manufacturing and application workers compared to the general population (Burgess et al., 2022; Dobraca et al., 2015; Trowbridge et al., 2020).

The United States National Academies of Sciences, Engineering, and Medicine (NASEM) recently provided clinical guidance for follow up with patients exposed to the following PFAS (NASEM PFAS): PFOA, PFNA, PFDA, PFUnDA, PFHxS, PFOS and N-methylperfluro-1-octanesulfonamido-acetic acid (MeFOSSA). The consensus report recommends that individuals with summed serum concentrations of NASEM PFAS ≥ 20 ng/ml should undergo clinical follow-up for thyroid function (age 18 or older); kidney function and cancer (age 45 or older); and ulcerative colitis and testicular cancer (age 15 or older)((NASEM, 2021). Individuals with the sum of the same PFAS ≥ 2 ng/ml should be prioritized for screening for dyslipidemia, hypertensive disorders of pregnancy, and breast cancer (at specified intervals). But interventions to reduce PFAS levels in the body are limited.

Biomonitoring efforts over the past decade have focused on assessing exposure to long-chain (perfluorosulfonic acids with 6 or more C atoms and perfluorocarboxylic acids with 7 or more C atoms), legacy PFAS such as the NASEM PFAS. Many legacy PFAS have been phased out of production in most parts of the world, and studies of general populations in North America show that body burdens of some long-chain PFAS are decreasing (Cakmak et al., 2022; Sonnenberg et al., 2023). However, analyses of environmental samples (e.g., water and wastewater) are revealing increasing trends for short-chain PFAS (C4-C7) such as perfluorobutanoic acid (PFBA), perfluoropentanoic acid (PFPeA), perfluorohexanoic acid (PFHxA) and perflurobutanesulfonic acid (PFBS) (Gewurtz et al., 2024). Trends in PFAS production and use also have started to be reflected in human serum PFAS concentrations worldwide, with serum concentrations of replacements for PFOA and PFOS increasing (Fan et al., 2022). Focusing on studies conducted in North America, the Canadian Health Measures Survey (CHMS) of the general Canadian population began analyzing short-chain PFAS (PFBA, PFHxA and PFBS) in Cycle 2 (2007-2009) but did not detect these PFAS in serum until sampling in Cycle 5 (2015-216) (Health Canada, 2021). The increase in detection frequency in Cycle 5 versus Cycle 2 may be explained in part by the lower limits of detection in Cycle 5, however. PFBA, PFHxA and PFBS were not detected in serum of First Nation populations in samples collected in 2016-17 (Aker et al., 2023; Caron-Beaudoin et al., 2019; Garcia-Barrios et al., 2021). In the 2013-2014 cycle of the United States National Health and Nutrition Examination Survey, small portions of the general population were found to have measurable levels of serum PFBS and PFHpA (0.6% and 12.6%, respectively) (Calafat et al., 2019). In 2020, however, short-chain PFAS and ultra-short-chain PFAS (C<4) were frequently detected in serum in samples collected in the United States, possibly stemming from indoor sources (Zheng et al., 2023). Exposure to these shorter-chained PFAS in other populations and their potential health risks are not well understood.

Here, we take advantage of serum samples collected as part of a clinical trial in 2019-2020 testing the efficacy of a dietary fiber intervention to reduce serum cholesterol, to investigate PFAS body burden of long-chain, short-chain and alternative PFAS in a Canadian population. In addition, we used clinical data on liver function biomarkers and serum lipid concentrations to examine associations between PFAS and these clinically relevant biomarkers. Last, given that bile acids and PFAS are biochemically similar as amphiphilic compounds, we examined whether the fiber intervention, which was designed to reduce cholesterol by trapping bile acids in the gut lumen, might reduce PFAS concentrations in serum.

## 2. Methods

### 2.1. Human serum samples

Serum samples were obtained from a previously conducted clinical trial (NCT03911427) that was conducted with the goal of reducing cholesterol through oat β-glucan (OBG) supplementation in adults 18-65y with elevated low density lipoprotein cholesterol (LDL-C) at baseline. Participants in the original study (224 in total) were recruited from Toronto, Ontario, Canada and the surrounding area from April 2019-February 2020 (Wolever et al., 2021). In the randomized, double-blind, placebo-controlled, parallel-arm design clinical trial, participants consumed sachets of OBG (1 g β-glucan, 1.9g fiber per sachet; intervention group) or brown rice powder (0 g β-glucan, 0.3g fiber per sachet; control group) mixed with 8 oz water three times per day separated by ≥3 h and preferably immediately before or within 10 min of each main meal (breakfast, lunch, and dinner)(Wolever et al., 2021). Detailed subject recruitment, study protocols and Institutional Review Board compliance for human subjects research are described elsewhere (Wolever et al., 2021). Fasting serum samples were collected at baseline and after 4 weeks of intervention (Wolever et al., 2021). Deidentified serum samples were transported on temperature monitored dry ice and blinded before PFAS analyses. Serum samples from the participants were analyzed for seventeen PFAS at baseline and after 4 weeks on the intervention. As determined by the Boston University Institutional Review Board, secondary analysis of previously collected, deidentified samples described herein does not constitute human subjects research.

Based on serum PFAS levels observed in the general North American (Canada and US) population, males have been reported to have greater concentrations of total conventional PFAS than females. (Health Canada, 2021; Lorber et al., 2015; Taylor et al., 2014; Wong et al., 2014) Using this information, we selected serum samples from male participants and restricted the sample pool to only those males who followed protocol (per protocol, PP) according to Wolever *et al*. 2021. Of the 224 original study participants, 74 were male, among which 72 were identified as “per protocol “ (PP; 42 PP in the OBG intervention group; 30 PP in the rice control group).

### 2.2 Biomarker analyses

Fasting serum samples were analyzed for total- and high density lipoprotein cholesterol (HDL-C), triglycerides, calculated LDL-C, aspartate aminotransaminase (AST), alanine aminotransferase (ALT) and gamma-glutamyl transferase (GGT) as described in the original study (Wolever et al., 2021).

### 2.3 PFAS serum analyses

The analytical method targeted 17 PFAS consisting of PFBuA, PFHxA, PFHxS, PFHpA, PFOA, PFOS, perfluorooctanesulfonamide (PFOSA), PFNA, PFDA, PFUdA, GenX, 1H,1H,2H,2H-Perflurooctanesulfonate (6:2 FTS), MeFOSSA, N-ethylperfluro-1-octanesulfonamidoacetic acid (Et-FOSAA), perfluoropentylsulfonate (L-PFPeS), PFDoA, and perfluro-4ethylcyclohexanesulfonate (PFECHS). Information on materials and samples preparation are provided in the Supplemental Material.

Serum samples (200 μL) were spiked with the internal standard (IS) cocktail (nominal 1-10 ng/ml each; IS mix cocktail obtained from Wellington Labs LLC, Wilmington, DE), followed by the addition of 1 ml of 1% acetic acid in distilled water to denature protein and centrifugation at 12500× G for 5 min. The supernatant was loaded into a prewashed and preconditioned solid phase extraction cartridge (Strata X-AW 33µm; 6mg/3ml; Phenomenex Inc, Torrance CA) for sample cleanup and concentration. The final eluate containing PFAS (2×1ml 1% ammonium hydroxide in methanol) was evaporated to dryness under a gentle stream of nitrogen, reconstituted in 200 μL methanol, and analyzed for PFAS.

PFAS were identified and quantified by liquid chromatography – negative electrospray ionization - tandem-mass spectrometry (LC-ESI^-^ - MS/MS) in an Applied Biosystems API4000 triple quadrupole mass spectrometer using the isotope dilution method. Chromatographic separation was accomplished on a Shimadzu LC20 series stack using a Luna C18, 3µm, 100×4.6mm analytical column (Phenomenex). Mobile phases were 10mM ammonium acetate in deionized water (A) and 10mM ammonium acetate in methanol (B). Chromatographic gradient was 10% B the first min, to 65% B at 2 min, to 99% B at 15 min, hold 99% B to 20 min, followed by 5 min post-column equilibration. Sample injection volume was 10µL. Background PFAS contamination was eliminated using an online delay column (Phenomenex Luna C18, 50x4.6 mm, 3µm). A diverter valve was used (VICI, Valco Instrument Co. Inc.) to divert the front and back-end of the chromatographic run to waste.

The scheduled MRM mode (**Table S1**) was used for data acquisition of the target set of PFAS. Twenty percent of samples were analyzed as true blind duplicates. Analyte recovery was 95-102% at serum concentrations of 0.5-5 ng/ml (<7% relative standard deviation), with calibration curve coefficients of R >0.999 for all analytes. Limits of detection ranged from 1 to 50 pg/ml, equivalent to 10-500 fg injection on the column. Tested and certified PFAS-free labware was used throughout the chain of sample acquisition, processing, and analysis. Quality control included laboratory blanks, blind sample duplicates, random blanks to check for carryover or cross-contamination, and a well-characterized aqueous film forming foam sample.

### 2.4 Statistical analyses

Demographic data were examined for underlying distributions using the Shapiro-Wilks statistical text, and ln-transformed values were used for subsequent statistical analysis (unpaired t-tests) performed with Prism 10.2.3 (GraphPad Inc. Boston, MA). Concentrations of individual PFAS in serum samples were examined for underlying distributions using the Shapiro-Wilks statistical text, and ln-transformed serum values were used for subsequent statistical analysis performed with SAS 9.4 (SAS Institute Inc. Cary, NC). Descriptive statistics including geometric means (GM) and geometric standard deviations (GSD) were calculated for 11 individual PFAS detected at baseline and after 4 weeks of intervention in 70% or more of participants, stratified by intervention (OBG vs. rice group). The GM ratios of serum concentrations between baseline and week 4 of intervention were calculated as the difference in values of the paired data for each individual PFAS. Paired t-tests on the ln-transformed data were conducted to examine the difference between PFAS serum concentrations at baseline and week 4 within each group. Multivariable linear regression was used to test associations between serum PFAS and clinical biomarkers (liver enzymes and cholesterol values) adjusting for age and BMI.

## 3. Results

### 3.1 Demographics and clinical biomarker values of study participants at baseline

The demographic data and clinical serum biomarker characteristics of the participants at baseline are presented in **Table 1**. These characteristics were similar between the two groups.

**Table 1:** Demographic and biomarker characteristics between men with oat β-glucan intervention versus control group^a^ at baseline.

|  | Total | OBG | Rice | p-value<sup>b</sup> |
| --- | --- | --- | --- | --- |
| **n** | 72 | 42 | 30 |  |
| **Age, y** | 41 ± 1 | 42 ± 11 | 41 ± 1 | 0.7200 |
| **Smoker (Y:N)** | 4:68 | 3:39 | 1:29 |  |
| **Wt (kg)** | 82.1 ± 1.2 | 82.6 ± 1.2 | 81.3 ± 1.2 | 0.7089 |
| **BMI (kg/m<sup>2</sup>)** | 27.2 ± 1.1 | 27.1 ± 1.2 | 27.3 ± 1.2 | 0.7721 |
| **Triglycerides (mmol/L)** | 1.48 ± 1.60 | 1.55 ± 1.58 | 1.38 ± 1.64 | 0.3013 |
| **T-Chol (mmol/L)** | 5.55 ± 1.12 | 5.59 ± 1.13 | 5.50 ± 1.11 | 0.5509 |
| **HDL-C (mmol/L)** | 1.17 ± 1.23 | 1.16 ± 1.26 | 1.19 ± 1.21 | 0.6328 |
| **LDL-C (mmol/L)** | 3.61 ± 1.14 | 3.62 ± 1.16 | 3.60 ± 1.12 | 0.8361 |
| **GGT (U/L)** | 24 ± 2 | 26 ± 2 | 27 ± 2 | 0.2166 |
| **ALT (U/L)** | 26 ± 2 | 25 ± 2 | 24 ± 1 | 0.4531 |
| **AST (U/L)** | 24 ± 1 | 23 ± 1 | 24 ± 1 | 0.3483 |

a. Values are geometric means ± geometric standard deviations or *n*.
b. Unpaired t-tests were performed on ln-transformed data.

### 3.2 PFAS distribution in serum at baseline

In serum samples collected at baseline, 11 of 17 PFAS were detected in >70% of participants (GenX, PFBuA, PFHpA, PFHxA, PFHxS, PFOA, PFOS, PFOSA PFNA, PFDA, PFUdA) while five of them (6:2 FTS, Me-FOSAA, Et-FOSAA, L-PFPeS, PFDoA, PFECHS), were below the method limit of detection (LOD, **Table S3**). Of the PFAS detected above the LOD, PFBuA, PFHxA, PFOA, PFHxS, PFOS and PFOSA were quantified in 100% of the samples. Five PFAS (GenX, PFHpA, PFNA, PFDA, PFUdA) were measured in 71-97% of samples (**Table 2**). The highest PFAS concentrations measured at baseline correspond to PFOSA (GM=7.1; GSD=1.8 ng/ml), PFOS (4.2; 1.9 ng/ml), PFOA (1.8;1.6 ng/ml) and PFHxS (1.3; 2.4 ng/ml) and the lowest levels correspond to PFDA and PFUnDA (**Table 2**).

**Table 2:** Descriptive statistics (geometric mean [geometric SD]) of serum concentrations of individual PFAS and their sum (ng/ml serum).

| PFAS | CHMS Cycle 6<br>Geometric<br>Mean (range,<br>ng/ml serum)<sup>c</sup> | Current Study |  |  |
| --- | --- | --- | --- | --- |
|  |  | LOD | %<br>Detection<br>at<br>Baseline<br>(n=72) | GM at Baseline<br>(ng/ml serum) |
| **PFBuA** | <LOD (0.075)<sup>d</sup> | <0.007 | 100 | 0.77 (1.55) |
| **PFHxA** | <LOD (0.084)<sup>d</sup> | <0.0015 | 100 | 0.59 (2.32) |
| **PFHpA** | NA | <0.002 | 90 | 0.44 (4.08) |
| **PFOA** | 1.4 (1.2-1.6) | <0.002 | 100 | 1.79 (1.69) |
| **PFNA** | 0.48 (0.44–0.53) | <0.001 | 97 | 0.84 (2.54) |
| **PFDA** | 0.13 (0.11–0.15) | <0.0015 | 88 | 0.11 (2.92) |
| **PFUdA** | 0.38 (0.30–0.46)<sup>e</sup> | <0.005 | 71 | 0.16 (4.11) |
| **PFHxS** | 1.2 (1.0–1.4) | <0.017 | 100 | 1.27 (2.42) |
| **PFOS** | 3.6 (3.3–4.0) | <0.007 | 100 | 4.16 (1.88) |
| **GenX<sup>a</sup>** | NA | <0.030 | 71 | 0.45 (8.66) |
| **PFOSA** | NA | <0.003 | 100 | 7.06 (1.79) |
| **Total**<br>**PFAS** |  |  |  | 21.51 (1.72) |
| **NASEM**<br>**PFAS<sup>b</sup>** |  |  |  | 9.19 (1.63) |

- Two samples had GenX concentrations 57 and 62 ng/ml (2-fold higher than the next highest concentration). 75<sup>th</sup> percentile = 1.25 ng/ml. 95<sup>th</sup> percentile = 16.76 ng/ml. - NASEM PFAS include PFOA, PFNA, PFDA, PFUnDA, PFHxS, PFOS. Me-FOSAA concentrations were below the LOD. - Canadian Health Measures Survey (CHMS) study (2018-2019). Measured in males 12-79 years of age (PFBuA, PFHxA, PFNA, PFDA, PFUdA) or 20-79 years of age (PFOA, PFHxS, PFOS). Health Canada, 2021. - LOD (limit of detection) is provided in parenthesis. - 90<sup>th</sup> percentile. - NA, Not analyzed; GM – Geometric mean.

We compared the serum PFAS concentrations of participants from Ontario, Canada to those observed in cycle 6 of CHMS (2018-2019) (Health Canada, 2021). Compared to concentrations reported in CHMS, serum concentrations of PFBuA and PFHxA were at least 7-10x higher (**Table 2**). All other PFAS had similar concentrations in both studies (**Table 2**).

### 3.3 Testing for associations of liver damage biomarkers with PFAS serum levels

Liver is a major target organ of PFAS, with PFAS inducing hepatomegaly and lipid accumulation in rodent models (e.g., (Das et al., 2017)). Therefore, we tested associations between PFAS serum levels and serum markers of liver damage GGT, AST and ALT. A number of PFAS, including PFBuA, PFHxA, PFDA and PFOSA, were associated with higher GGT concentrations, while PFHxA and PFHxS were associated with higher ALT concentrations (**Table 3, Table S2**). We observed no associations between PFAS analyzed in serum and AST concentrations.

**Table 3:** Association between liver function biomarkers and PFAS exposure (adjusted for age and BMI).

| PFAS | GGT |  | AST |  | ALT |  |
| --- | --- | --- | --- | --- | --- | --- |
| | $\beta^a$ | p-value<sup>b</sup> | $\beta$ | p-value | $\beta$ | p-value |
| PFBuA | 0.32 | **0.009** | -0.036 | 0.649 | 0.182 | 0.152 |
| PFHxA | 0.121 | **0.054** | 0.054 | 0.171 | 0.125 | **0.051** |
| PFHpA | -0.022 | 0.558 | 0.008 | 0.735 | 0.022 | 0.579 |
| PFOA | 0.12 | 0.281 | 0.09 | 0.193 | 0.111 | 0.331 |
| PFNA | 0.064 | 0.279 | -0.007 | 0.853 | -0.027 | 0.654 |
| PFDA | 0.094 | **0.062** | -0.024 | 0.445 | -0.038 | 0.463 |
| PFUDa | -0.006 | 0.88 | -0.03 | 0.217 | -0.059 | 0.146 |
| PFHxS | -0.033 | 0.605 | 0.044 | 0.259 | 0.151 | **0.017** |
| PFOS | 0.019 | 0.841 | -0.011 | 0.849 | -0.001 | 0.99 |
| GenX | -0.021 | 0.43 | -0.019 | 0.245 | -0.045 | 0.087 |
| PFOSA | 0.185 | **0.059** | 0.068 | 0.266 | 0.148 | 0.141 |
| Total PFAS | 0.076 | 0.489 | 0.033 | 0.627 | 0.049 | 0.66 |
| NASEM PFAS<sup>c</sup> | 0.062 | 0.614 | 0.027 | 0.72 | 0.095 | 0.448 |

a. $\beta$ = Change in units of serum liver biomarker per ln-ng PFAS increase in serum.
b. Multivariable linear regression
c. NASEM PFAS include PFOA, PFNA, PFDA, PFUnDA, PFHxS, PFOS. Me-FOSAA concentrations were below the LOD.

### 3.3 Testing for associations of serum lipids with PFAS serum levels

Given the association of several PFAS with biomarkers of liver damage, that the liver is the central hub of cholesterol metabolism (Dietschy et al., 1993), and that increased total cholesterol and non-HDL-C are well-supported health endpoints linked to PFAS exposure in humans (Schlezinger and Gokce, 2024), we assessed the relationship between PFAS and blood lipid levels. Only PFHxA and PFHpA were significantly associated with serum cholesterol concentrations, and they were associated with higher concentrations of HDL-C (**Table 4, Table S3**).

**Table 4:**
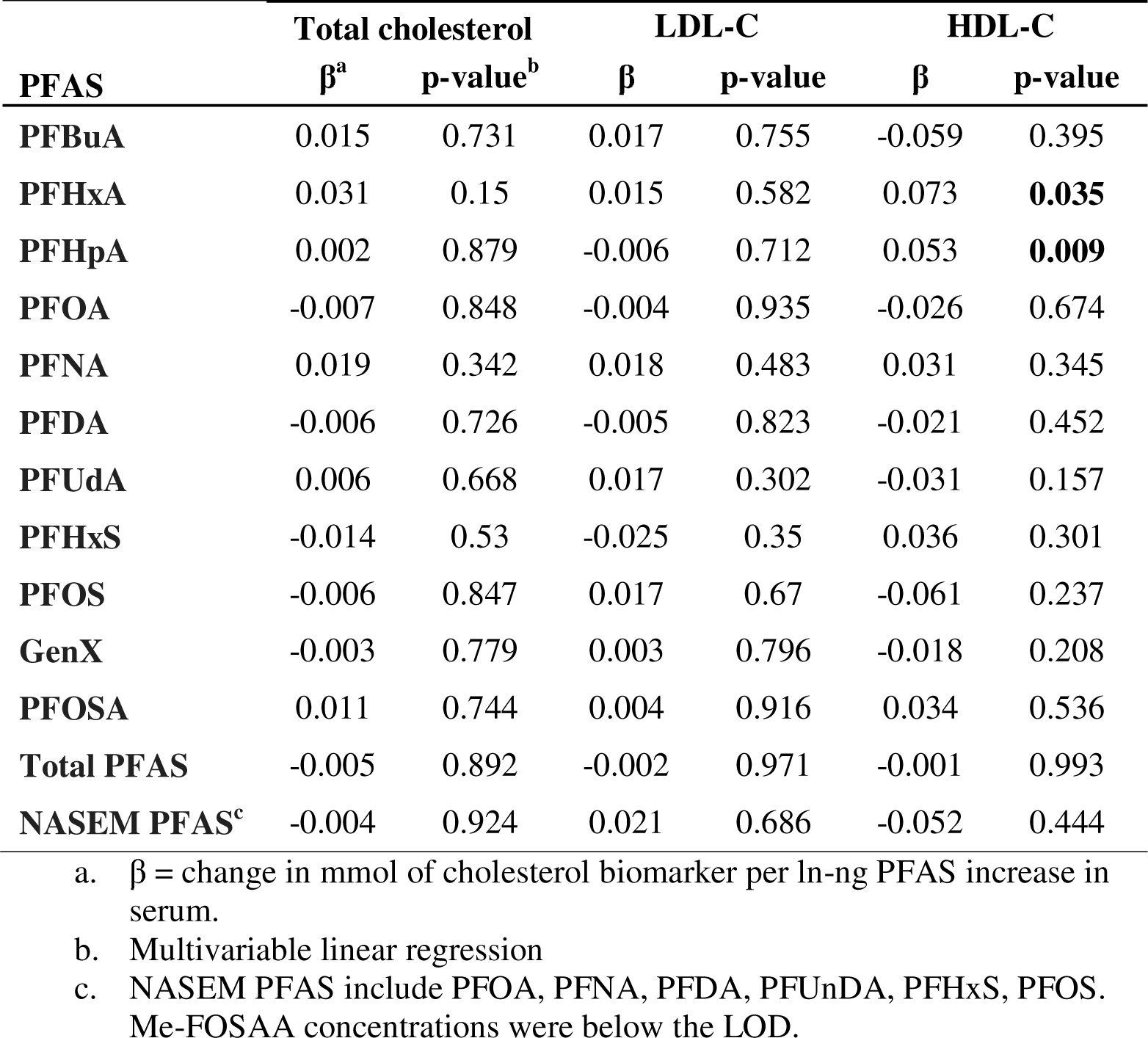
Associations between measures of serum cholesterol and PFAS exposure (adjusted for age and BMI).

| PFAS | Total cholesterol |  | LDL-C |  | HDL-C |  |
| --- | --- | --- | --- | --- | --- | --- |
| | $\beta^a$ | p-value<sup>b</sup> | $\beta$ | p-value | $\beta$ | p-value |
| PFBuA | 0.015 | 0.731 | 0.017 | 0.755 | -0.059 | 0.395 |
| PFHxA | 0.031 | 0.15 | 0.015 | 0.582 | 0.073 | **0.035** |
| PFHpA | 0.002 | 0.879 | -0.006 | 0.712 | 0.053 | **0.009** |
| PFOA | -0.007 | 0.848 | -0.004 | 0.935 | -0.026 | 0.674 |
| PFNA | 0.019 | 0.342 | 0.018 | 0.483 | 0.031 | 0.345 |
| PFDA | -0.006 | 0.726 | -0.005 | 0.823 | -0.021 | 0.452 |
| PFUnDA | 0.006 | 0.668 | 0.017 | 0.302 | -0.031 | 0.157 |
| PFHxS | -0.014 | 0.53 | -0.025 | 0.35 | 0.036 | 0.301 |
| PFOS | -0.006 | 0.847 | 0.017 | 0.67 | -0.061 | 0.237 |
| GenX | -0.003 | 0.779 | 0.003 | 0.796 | -0.018 | 0.208 |
| PFOSA | 0.011 | 0.744 | 0.004 | 0.916 | 0.034 | 0.536 |
| Total PFAS | -0.005 | 0.892 | -0.002 | 0.971 | -0.001 | 0.993 |
| NASEM PFAS<sup>c</sup> | -0.004 | 0.924 | 0.021 | 0.686 | -0.052 | 0.444 |

a. $\beta$ = change in mmol of cholesterol biomarker per ln-ng PFAS increase in serum.
b. Multivariable linear regression
c. NASEM PFAS include PFOA, PFNA, PFDA, PFUnDA, PFHxS, PFOS. Me-FOSAA concentrations were below the LOD.

### 3.4 Assessing the impact of a dietary fiber intervention on PFAS serum levels

The original study, from which the samples were collected, was designed to test the efficacy of an oat β-glucan beverage in reducing serum cholesterol levels (Wolever et al., 2021). Gel-forming fibers, such as β-glucan, reduce cholesterol by trapping bile acids in the gut lumen, thereby enhancing their excretion (Yu et al., 2022). Given that PFAS share biochemical characteristics with bile acids, we examined the data to determine if PFAS serum concentrations were reduced to a greater extent in participants that consumed the oat β-glucan beverage versus a rice beverage. Total PFAS concentrations decreased in both groups; however, there was no difference in the magnitude of decrease between the OBG and Rice groups (**Table 5**). Of the individual PFAS, PFBuA, PFHxA and PFHpA decreased significantly in concentration in both groups over the 4-week study period (**Table 5**). However, NASEM PFAS significantly decreased in concentration only in the OBG group over the study period, likely driven by a significant decrease in PFHxS (**Table 5).**

**Table 5:** Effect of fiber beverage intervention on serum PFAS concentrations.

| PFAS | OBG |  |  |  | Rice |  |  |  |
| --- | --- | --- | --- | --- | --- | --- | --- | --- |
|  | *Baseline* | *4 weeks* | *GM Ratio<sup>a</sup>* | *p-value<sup>b</sup>* | *Baseline* | *4 weeks* | *GM Ratio<sup>a</sup>* | *p-value<sup>b</sup>* |
|  | GM (GSD) | GM (GSD) | B/W4 |  | GM (GSD) | GM (GSD) | B/W4 |  |
| **PFBuA** | 0.80 (1.6) | 0.66 (1.6) | 1.2 | **<0.01** | 0.73 (1.5) | 0.58 (1.6) | 1.3 | **0.01** |
| **PFHxA** | 0.59 (2.4) | 0.51 (2.2) | 1.2 | **0.06** | 0.58 (2.2) | 0.45 (1.9) | 1.3 | **0.02** |
| **PFHpA** | 0.37 (4.5) | 0.30 (4.4) | 1.2 | **<0.01** | 0.56 (3.4) | 0.38 (2.9) | 1.5 | **0.01** |
| **PFOA** | 1.74 (1.7) | 1.61 (1.8) | 1.1 | 0.14 | 1.87 (1.6) | 1.85 (1.8) | 1 | 0.8 |
| **PFNA** | 0.70 (3.1) | 0.66 (3.1) | 1.1 | 0.14 | 1.07 (1.7) | 1.07 (1.7) | 1 | 0.98 |
| **PFDA** | 0.10 (3.2) | 0.10 (3.2) | 1 | 0.84 | 0.13 (2.5) | 0.14 (2.5) | 0.9 | 0.4 |
| **PFUnA** | 0.13 (4.4) | 0.13 (4.3) | 1 | 0.8 | 0.20 (3.7) | 0.20 (3.7) | 1 | 0.89 |
| **PFHxS** | 1.09 (2.6) | 1.00 (2.5) | 1.1 | **0.07** | 1.56 (2.2) | 1.53 (1.8) | 1 | 0.88 |
| **PFOS** | 3.91 (1.8) | 3.59 (2.0) | 1.1 | 0.12 | 4.50 (2.0) | 4.31 (2.1) | 1 | 0.35 |
| **GenX** | 0.29 (6.4) | 0.28 (6.7) | 1 | 0.77 | 0.83 (11.0) | 0.66 (8.1) | 1.3 | 0.29 |
| **PFOSA** | 6.91 (1.8) | 6.55 (1.8) | 1.1 | 0.5 | 7.27 (1.8) | 7.35 (1.8) | 1 | 0.78 |
| **Total PFAS** | 19.2 (1.5) | 17.7 (1.6) | 1.1 | **0.08** | 24.9 (1.9) | 21.6 (1.6) | 1.2 | **0.05** |
| **NASEM PFAS<sup>a</sup>** | 8.6 (1.6) | 7.9 (1.7) | 1.1 | **0.03** | 10.1 (1.6) | 9.7 (1.7) | 1 | 0.27 |

- GM ratios were calculated as the difference in ln-serum concentrations at baseline (B) and week 4 (W4) of the intervention of the paired data for each specific PFAS, NASEM PFAS and total sum. - GM concentrations are reported in ng/ml - Paired t-tests were performed on ln-transformed data to compare PFAS concentrations at baseline vs. week 4. - NASEM PFAS include PFOA, PFNA, PFDA, PFUnDA, PFHxS, PFOS. Me-FOSAA concentrations were below the LOD in all samples.

## 4. Discussion

Targeting a broader and more diverse list of PFAS analytes than in most previously published studies, we aimed to characterize long-chain, short-chain and alternative PFAS in previously collected serum from Canadian participants from a published clinical trial. Also, using the available liver and lipid homeostasis biomarker data, we testes the associations of PFAS with these biomarkers. Finally, we took the opportunity to pilot test a novel hypothesis, that consumption of gel-forming dietary fibers can decrease PFAS body burdens in people.

Serum concentrations of long-chain PFAS in this study were largely typical of those found in the general population of Canada. Overall, the participants in this study had serum PFAS concentrations consistent with typical food/water borne exposure and not with occupational exposures or exposures associated with living in a community heavily contaminated with PFAS (Burgess et al., 2022; Dobraca et al., 2015; Trowbridge et al., 2020). Regardless, all of the participants had NASEM PFAS serum concentrations ≥ 2 ng/ml, the exposure level at which NASEM clinical guidance recommends decreasing exposure and further medical follow-up (screening for dyslipidemia, hypertensive disorders of pregnancy and breast cancers). Seven percent of participants had NASEM PFAS serum concentrations ≥ 20 ng/ml. Per NASEM guidelines, these individuals should undergo clinical follow-up for thyroid function (age 18 or older); kidney function and cancer (age 45 or older); and ulcerative colitis and testicular cancer (age 15 or older)((NASEM, 2021).

Long-chain precursors (e.g., PFOSA, a precursor to PFOS), short-chain PFAS and alternative PFAS (e.g., GenX) were commonly detected in baseline serum samples. One previous study attempted to quantify PFOSA in Canadians in samples taken in 2002, yet all samples were below the LOD of 1.5 ng/ml (Kubwabo et al., 2004). In this study, PFOSA was detectable in all baseline samples and had the highest geometric mean serum concentration of any PFAS measured (7.1 ng/ml). This is the first study to report GenX exposure in a Canadian population; we detected GenX in 71% of baseline serum samples. Of note, we detected short-chain PFBuA and PFHxA in nearly 100% of baseline serum samples, not previously detectable in CHMS Cycle 6. We detected PFHpA in 90% of serum samples, a higher frequency of detection than was observed in samples of pregnant Canadians taken in 2009-12 (67% (Reardon et al., 2023)) or 2004-2005 (<5% (Monroy et al., 2008)). While lower LODs in this study may play a role in the higher detection frequencies, they may also reflect increasing production and use of these PFAS (Zheng et al., 2023).

Several adverse health effects have been linked to PFAS and are well-supported by epidemiological data in humans (ATSDR, 2021). Here, we were specifically interested in testing the associations of PFAS serum concentrations with biomarkers of liver toxicity and dyslipidemia, as they are likely mechanistically linked (Dietschy et al., 1993). A recent meta-analysis reported associations between PFOA, PFNA, PFOS, and PFHxS and higher serum ALT levels, with PFOA also being associated with higher GGT levels (Costello et al., 2022). Here, we observed that PFHxS also was associated with higher serum ALT and multiple PFAS (PFBuA, PFHxA, PFDA, and PFOSA) were associated with higher GGT concentrations. This is in line with another recent study of Canadian participants that showed that GGT was the liver function biomarker most commonly associated with multiple PFAS (Cakmak et al., 2022).

We expected to observe positive associations between total cholesterol and LDL-C measures and PFAS serum concentrations, as the epidemiological data supporting these associations are robust (Schlezinger and Gokce, 2024) and they have been reported in Canadians (Cakmak et al., 2022). However, the only associations that reached significance were for higher levels of HDL-c associated with PFHpA and PFHxA, possibly due to the relatively small sample size in this study compared to Cakmak et al. 2022 (72 participants vs. 6768 participants). There are multiple reasons why we detected associations between PFAS serum concentrations and biomarkers of liver injury and not serum cholesterol. Since perturbation of lipid metabolism is likely to occur downstream of liver injury, changes in serum cholesterol may not have had sufficient time to develop. Additionally, differences in precision of the analyses may allow for greater sensitivity to detect differences in biomarkers of liver injury than changes in serum cholesterol.

Many PFAS are ionized at physiological pH and thus are transported from the gut contents into the body by cells that line the intestines (Zhao et al., 2017), leading to efficient initial absorption following ingestion of PFAS (Pizzurro et al., 2019) and reabsorption after excretion of PFAS in bile (Cao et al., 2022). The cycle of excretion in bile and reuptake by gut lining cells via enterohepatic recirculation is an important factor contributing to the years’ long half-life of conventional PFAS in humans (Cao et al., 2022). As such, reducing enterohepatic recirculation is anticipated to reduce body burdens of PFAS. Numerous studies have shown that consumption of gel-forming dietary fibers (e.g., β-glucan found in oats and barley) (Yu et al., 2022) leads to reductions in LDL-C in the blood because these fibers “trap” bile acids, which are formed from cholesterol in the liver. Trapping of bile acids in the gut lumen thereby decreases the potential for enterohepatic recirculation and enhances their fecal excretion (Silva et al., 2021). Consistent with these pharmacokinetic properties of ionizable PFAS, decreased serum PFAS concentrations have been associated with increased fiber intake (Dzierlenga et al., 2021).

Taking advantage of the original clinical study design, which was designed to test the ability of an oat β-glucan beverage to reduce serum cholesterol concentrations, a pilot analysis of the effect of consumption of a gel-forming fiber on PFAS serum concentrations was conducted. Total Σ11 PFAS concentrations decreased overall in both study groups. This may reflect the fact that short chain PFAS have half-lives on the order of In addition, significant reductions in some PFAS in both groups may reflect shorter human half-lives of short-chain PFAS (e.g., 62 days for PFHpA (Xu et al., 2020). Interestingly, only the oat β-glucan group saw significant reductions in the NASEM PFAS concentration, which appeared to have been driven by reductions in PFHxS. It is not necessarily surprising that differences in effect across PFAS types are seen, given that the ratio of urinary to biliary PFAS excretion depends upon PFAS structure (Fujii et al., 2015).

There are several likely reasons why a stronger effect of the intervention was not observed. Samples used in the current study were from a previously conducted study that was not designed as an intervention to reduce PFAS; thus, there was no information collected on potential sources of PFAS exposure prior to or during the study and no attempt to control for differences in ongoing exposures between intervention and control groups. Further, the time of the intervention was short, only 4 weeks. Many conventional PFAS have half-lives on the order of 2-7 years (Bartell, 2012; Ji et al., 2021; Li et al., 2022; Russell et al., 2015; Zhang et al., 2013), thus a one-month intervention may be insufficient to strongly influence serum PFAS concentrations with ongoing exposure. Along these lines, a recent cholestyramine intervention study in which the ability of the anion resin to trap PFAS in the gut lumen was tested, showed that a 12-week intervention induced notable reductions in serum concentrations of PFAS (Møller et al., 2024). A meta-analysis of barley β-glucan showed that consumption of a minimum of 6.5-6.9g β-glucan per day was necessary to reduce serum cholesterol (Ho et al., 2016), which is higher than the 3g oat β-glucan consumed daily in the current study. Thus, a higher consumption of β-glucan is possible and may also improve PFAS reductions.

## 5. Conclusions

The types of PFAS to which humans are being exposed are evolving, yet the bulk of exposure assessment has focused on long-chain PFAS. As such, broader biomonitoring is needed to capture changing PFAS exposures. Here, we show that short-chain and alternative PFAS are measurable in Canadians, along with the PFOS-precursor PFOSA. Given that this is the first time some of these PFAS have been measured in Canadians, follow-up studies are needed to define exposure trends. For a subset of participants, we observed PFAS serum concentrations that exceeded NASEM guidelines of 20 ng/ml. Despite the limited sample size and cross-sectional nature of the biomarker analysis, we found positive associations between PFBuA and PFHxA and biomarkers of liver toxicity, suggesting potential health impacts for short-chained PFAS that warrant further investigation. Current clinical treatments to reduce PFAS body burden are minimal. Results from this pilot analysis suggest a potentially practical and feasible intervention that may reduce human body burdens for some PFAS. However, studies utilizing a larger sample with a broader range of serum concentrations, longer intervention period, and clinically relevant fiber intakes are needed to determine the efficacy of using gel-forming dietary fibers to increase PFAS excretion.

## 6. Abbreviations

6:2 FTS: 1H,1H,2H,2H-Perflurooctanesulfonate
CHMS: Canadian Health Measures Survey
Et-FOSAA: N-Ethylperfluro-1-octanesulfonamidoacetic acid
GenX: Undecafluoro-2-methyl-3-oxahexanoic acid
HDL-C: High density lipoprotein cholesterol
LDL-C: Low density lipoprotein cholesterol
LOD: Limit of detection
L-PFPeS: Perfluoropentylsulfonate
Me-FOSAA: N-Methylperfluro-1-octanesulfonamidoacetic acid
NASEM: United States National Academies of Sciences, Engineering and Medicine
OBG: Oat β-glucan
PFAS: Per and polyfluoroalkyl substances
PFBS: Perfluorobutane sulfonic acid
PFBuA: Perfluorobutanonic Acid
PFCA: Perfluoroalkyl carboxylic acid
PFDA: Perfluorodecanoic acid
PFDoA: Perfluorododecanoic acid
PFECHS: Perfluro-4-ethylcyclohexanesulfonate
PFHpA: Perfluoroheptanoic acid
PFHxA: Perfluorohexanoic Acid
PFHxS: Perfluorohexane sulfonic acid
PFNA: Perfluorononanoic acid
PFOA: Perfluorooctanoic acid
PFOS: Perfluorooctane sulfonic acid
PFOSA: Perfluorooctanesulfonamide
PFSA: Perfluoroalkyl sulfonic acid
PFUdA: Perfluoroundecanoic acid
PP: Per protocol

## Data Availability

All data produced in the present study are available upon reasonable request to the authors.

