## Supplemental Material for "Per- and Poly-fluoroalkyl Substances (PFAS) in Circulation in a Canadian Population: Their Association with Serum Liver Enzyme Biomarkers and Piloting a Novel Method to Reduce Serum PFAS"

**Table of Contents**

Page 2: Materials and methods for PFAS analyses

Page 3: Table S1. Multiple Reactions Monitoring (MRM) of PFAS

Page 4: Table S2. List of PFAS internal standards associated with PFAS

Page 5: Table S3. PFAS limits of detection.

Page 6: Table S3. Association between liver function biomarkers and PFAS exposure (Unadjusted).

Page 7: Table S4. Associations between measures of serum cholesterol and PFAS exposure (Unadjusted).

**Materials and Methods for PFAS Analyses**

**Materials**

High purity liquid chromatography grade solvents, methanol (Cat# BDH85800.400) and acetonitrile (Cat# BDH83639.400) were purchased from VWR International (Radnor, PA). High purity deionized water, >18MΩ, was obtained from a Milli-Q Direct 8 unit (Millipore Sigma). LC-MS grade ammonium acetate was purchased from Sigma Aldrich (Chicago, IL). HPLC analytical column (Luna^®^ Omega PS C18, 3µm, 100 Å, 100 × 4.6 mm, Part no. 00D-4758-E0) and guard column cartridge (SecurityGuard^TM^ PS C18, 4 × 3.0 mm, Part No. AJ0-7606) were obtained from Phenomenex (Torrance, CA). A PFAS delay column (Ascentis^®^ Express 160 Å FAS Delay, 2.7µm, 50 × 4.6mm, Cat#53573-U) was purchased from Supelco^®^, Millipore Sigma (Atlanta, GA). Solid phase extraction cartridge (Strata^TM^ X-AW 33µm polymeric weak anion; 60mg/3mL, Cat#8B-S038-UBL) were obtained from Phenomenex Inc (Torrance CA). Polypropylene LC-MS vials (SureSTART Polypropylene Microvials, Cat#6ESV9-1PP) were purchased from Fisher Scientific Company (Hanover Park, IL). A PFAS standard mixture (PFAC-MXF, -MXG, -MXH, -MXI, -MXJ), which contained sixteen PFAS standards quantified in this study [PFBuA, PFHxA, PFHxS, PFHpA, PFOA, PFOS, PFOSA, PFNA, PFDA, PFUdA, 6:2 FTS, Me-FOSAA, Et-FOSAA, L-PFPeS, PFDoA, and PFECHS] were purchased from Wellington Labs LLC (Wilmington, DE); undecafluoro-2-methyl-3-oxahexanoic Acid (GenX) was obtained from Synquest Laboratories (Alachua, FL). Isotopic-labelled PFAS stock solutions (MPFAC-HIF-ES and MPFAC-HIF-IS) were also purchased from Wellington Labs LLC (Wilmington, DE) and used as indicated in Table S2.

**Table S1. Multiple Reactions Monitoring (MRM) of PFAS**

| # | **List of PFAS** | **CAS#** | **Abb.** | **MW** | **Molecular Ion (Q1)** | **Product Ion (Q3)** | **DP** | **EP** | **CE** | **CXP** |
| --- | --- | --- | --- | --- | --- | --- | --- | --- | --- | --- |
| 1 | Perfluoro-n-butanoic Acid | 375-22-4 | PFBuA | 214.06 | 213 | 169 | -32 | -10 | -13 | -10 |
|  |  |  |  |  |  | 101 | -32 | -10 | -29 | -16 |
| 2 | Perfluorohexanoic acid | 307-24-4 | PFHxA | 314.06 | 313 | 269 | -35 | -10 | -13 | -12 |
|  |  |  |  |  |  | 119 | -35 | -10 | -27 | -10 |
| 3 | Potassium perfluorohexanesulfonate | 3871-99-6 | PFHxS | 438.2 | 399 | 98.9 | -80 | -10 | -70 | -5 |
| 4 | Perfluoroheptanoic acid | 375-85-9 | PFHpA | 364.06 | 363 | 168.6 | -40 | -10 | -24 | -14 |
| 5 | Perfluorooctanoic acid | 335-67-1 | PFOA | 414 | 413 | 168.6 | -40 | -10 | -24 | -14 |
| 6 | Perfluorononanoic acid | 375-95-1 | PFNA | 464.07 | 463 | 419 | -40 | -10 | -15 | -10 |
|  |  |  |  |  |  | 169 | -40 | -10 | -27 | -8 |
| 7 | Perfluoro-1-octanesulfonamide | 754-91-6 | PFOSA | 499.1 | 498 | 80 | -75 | -10 | -99 | -10 |
|  |  |  |  |  |  | 98.7 | -75 | -10 | -72 | -10 |
| 8 | Perfluorooctanesulfonate | 1763-23-1 | PFOS | 500 | 499 | 98.8 | -80 | -10 | -60 | -10 |
| 9 | Perfluorodecanoic acid | 335-76-2 | PFDA | 514.08 | 513 | 469 | -45 | -10 | -15 | -10 |
|  |  |  |  |  |  | 269 | -45 | -10 | -25 | -10 |
| 10 | Perfluoroundecanoic acid | 2058-94-8 | PFUdA | 564.08 | 563 | 519 | -40 | -10 | -15 | -10 |
|  |  |  |  |  |  | 319 | -40 | -10 | -27 | -10 |
| 11 | Perfluorododecanoic acid | 307-55-1 | PFDoA | 614.08 | 613 | 569 | -45 | -10 | -17 | -13 |
|  |  |  |  |  |  | 293 | -45 | -10 | -14 | -10 |
| 12 | N-methylperfluro-octanesulfonamidoacetic acid | 2355-31-9 | Me-PFOSAA | 571.2 | 570 | 419 | -65 | -10 | -28 | -9 |
| 13 | N-ethylperfluro-octanesulfonamidoacetic acid | 2991-50-6 | Et-PFOSAA | 585.2 | 584 | 419 | -70 | -10 | -29 | -21 |
|  |  |  |  |  |  | 526 | -70 | -10 | -26 | -11 |
| 14 | Sodium 1H,1H,2H,2H-Perflurooctanesulfonate | 27619-94-9 | 6:2 FTS | 450 | 427 | 407 | -50 | -10 | -33 | -15 |
|  |  |  |  |  |  | 81 | -50 | -10 | -67 | -13 |
| 15 | Sodium perfluoro-1-pentanesulfonate | 630402-22-1 | LPFPeS | 372.08 | 349 | 80 | -75 | -10 | -66 | -7 |
|  |  |  |  |  |  | 99 | -75 | -10 | -48 | -7 |
| 16 | Potassium perfluro-4ethylcyclohexanesulfonate | 335-24-0 | PFECHS | 502.2 | 461 | 99 | -60 | -10 | -54 | -7 |
|  |  |  |  |  |  | 381 | -60 | -10 | -37 | -15 |
| 17 | Undecafluoro-2-methyl-3oxahexanoic Acid | 13252-13-6 | GenX | 330.05 | 329.2 | 285 | -25 | -10 | -8 | -7 |
|  |  |  |  |  |  | 168.8 | -25 | -10 | -18 | -8 |

**Table S2: List of PFAS Internal Standards associated with PFAS**

| # | PFAS Internal Standards | Abbreviation | Molecular Weight | Molecular Ion (Q1) | Product Ion (Q3) | Associated PFAS |
| --- | --- | --- | --- | --- | --- | --- |
| 1 | Perfluoro-n-13C3-butanoic Acid | M3PFBA | 217.01 | 216 | 172 | PFBuA, PFBS^*^, LPFPeS^*^ |
| 2 | Perfluoro-n-(1,2,13C2)-hexanoic acid | M5PFHxA | 319.02 | 318 | 273 | PFHxA, PFHxS^*^, GenX^*^ |
| 3 | Perfluoro-n-(1,2,3,4-13C4)-heptanoic acid | M4PFHpA | 368.03 | 367 | 322 | PFHpA, PFECHS^*^ |
| 4 | Perfluoro-n-13C8-octanoic acid | M8PFOA | 422 | 421 | 376 | PFOA |
| 5 | Perfluoro-n-13C9-nonanoic acid | M9PFNA | 520 | 519 | 474 | PFNA |
| 6 | Perfluoro-1-(13C8)-octanesulfonamide | M8FOSA | 507 | 506 | 78 | PFOSA |
| 7 | Perfluoro-1-(1,2,3,4-13C4)-octanesulfonate | MPFOS | 526.08 | 503 | 80 | PFOS |
| 8 | Perfluoro-n-(1,2-13C2)-decanoic acid | MPFDA | 516.07 | 515 | 470 | PFDA |
| 9 | Perfluoro-n-13C7-undecanoic acid | M7PFUdA | 571 | 570 | 525 | PFUdA |
| 10 | Perfluoro-n-13C2-dodecanoic acid | MPFDoA | 616 | 615 | 570 | PFDoA |
| 11 | N-methyl-d3-perfluro-octanesulfonamidoacetic acid | d3-MeFOSAA | 574.22 | 573 | 419 | Me-FOSAA |
| 12 | N-ethyl-d5-perfluro-octanesulfonamidoacetic acid | d5-EtPFOSAA | 590.27 | 589 | 419 | Et-PFOSAA |
| 13 | Sodium 1H,1H,2H,2H-(13C2)-Perflurooctanesulfonate | M2-6:2 FTS | 552 | 529 | 81 | 6:2 FTS |

^*^These PFAS do not have their own isotope labelled Internal Standard and a surrogate internal standard closest to the analyte retention time and molecular weight/structure was used as indicated in the table.

**Table S3: PFAS limits of detection (LOD).**

| Analyte | Lowest Detectable std concentration (ng/ml)* | S/N ratio | LOQ (ng/mL) | LOD (ng/ml) | R2 |
| --- | --- | --- | --- | --- | --- |
| PFBuA | 0.025 | 5.5 | 0.023 | 0.014 | 0.9995 |
| PFHxA | 0.0125 | 4.8 | 0.013 | 0.008 | 0.9998 |
| PFHxS | 0.0125 | 5.2 | 0.012 | 0.007 | 0.9998 |
| PFHpA | 0.0125 | 5 | 0.013 | 0.008 | 0.9998 |
| PFOA | 0.0125 | 4.6 | 0.014 | 0.008 | 0.9995 |
| PFNA | 0.0125 | 3.9 | 0.016 | 0.010 | 0.9997 |
| PFDA | 0.0125 | 5.01 | 0.012 | 0.007 | 0.9994 |
| PFUdA | 0.0125 | 3.3 | 0.019 | 0.011 | 0.9999 |
| PFDoA | 0.0125 | 4.2 | 0.015 | 0.009 | 0.9996 |
| PFOS | 0.025 | 6.5 | 0.019 | 0.012 | 1 |
| PFOSA | 0.025 | 4.4 | 0.028 | 0.017 | 1 |
| L-PFPeS | 0.0125 | 4.5 | 0.014 | 0.008 | 0.9999 |
| LFECHS | 0.025 | 8.5 | 0.015 | 0.009 | 0.9998 |
| 6:2 FTS | 0.01 | 4.1 | 0.012 | 0.007 | 0.9999 |
| Me-FOSAA | 0.05 | 3.5 | 0.071 | 0.043 | 0.9999 |
| Et-FOSAA | 0.125 | 5.5 | 0.114 | 0.068 | 1 |
| GenX | 0.025 | 5.2 | 0.024 | 0.014 | 0.9999 |
| NBP2 | 0.025 | 5.1 | 0.025 | 0.015 | 0.9999 |
| PFEtSA | 0.12 | 4.6 | 0.130 | 0.078 | 0.9997 |
| PFPrA | 0.5 | 4.8 | 0.521 | 0.313 | 0.9996 |
| PFPrS | 0.25 | 4.5 | 0.278 | 0.167 | 0.9995 |

***** 100 μl of serum was analyzed in a final total volume of 200 μL.

**Results**

**Table S3. Association between liver function biomarkers and PFAS exposure (Unadjusted).**

| **LB^a^ PFAS** | **GGT** |  |  | **AST** |  |  | **ALT** |  |
| --- | --- | --- | --- | --- | --- | --- | --- | --- |
|  | **β^b^** | **p-value^c^** |  | **β** | **p-value** |  | **β** | **p-value** |
| **PFBuA** | 0.25 | **0.04** |  | -0.05 | 0.55 |  | 0.12 | 0.33 |
| **PFHxA** | 0.12 | **0.07** |  | 0.05 | 0.19 |  | 0.12 | **0.09** |
| **PFHpA** | -0.02 | 0.57 |  | 0.01 | 0.80 |  | 0.02 | 0.69 |
| **PFOA** | 0.10 | 0.34 |  | 0.04 | 0.52 |  | 0.00 | 0.97 |
| **PFNA** | 0.07 | 0.25 |  | -0.02 | 0.66 |  | -0.05 | 0.41 |
| **PFDA** | 0.08 | **0.09** |  | -0.03 | 0.31 |  | -0.06 | 0.25 |
| **PFUdA** | -0.01 | 0.77 |  | -0.04 | 0.11 |  | -0.08 | **0.03** |
| **PFHxS** | -0.01 | 0.92 |  | 0.04 | 0.25 |  | 0.15 | **0.01** |
| **PFOS** | 0.01 | 0.93 |  | -0.04 | 0.44 |  | -0.09 | 0.31 |
| **GenX** | 0.17 | **0.06** |  | 0.03 | 0.55 |  | 0.07 | 0.49 |
| **PFOSA** | -0.02 | 0.39 |  | -0.02 | 0.12 |  | -0.06 | **0.02** |
| **Total PFAS** | 0.07 | 0.48 |  | -0.01 | 0.93 |  | -0.04 | 0.68 |
| **NASEM PFAS^d^** | 0.05 | 0.66 |  | -0.02 | 0.78 |  | -0.04 | 0.76 |

a. LB: Liver injury biomarker
b. β = Change in units of serum liver biomarker per ln-ng PFAS increase in serum.
c. Multivariable linear regression
d. NASEM PFAS include PFOA, PFNA, PFDA, PFUnDA, PFHxS, PFOS. Me-FOSAA concentrations were below the LOD.

**Table S4. Associations between measures of serum cholesterol and PFAS exposure (Unadjusted). β, mmol/μg PFAS in serum.**

| **CB^a^** **PFAS** | **Total cholesterol** | |  | **LDL** | |  | **HDL** | |
| --- | --- | --- | --- | --- | --- | --- | --- | --- |
|  | **β^b^** | **p-value^c^** |  | **β** | **p-value** |  | **β** | **p-value** |
| **PFBuA** | -0.008 | 0.854 |  | -0.007 | 0.891 |  | -0.052 | 0.453 |
| **PFHxA** | 0.030 | 0.180 |  | 0.013 | 0.625 |  | 0.076 | **0.034** |
| **PFHpA** | 0.004 | 0.783 |  | -0.004 | 0.800 |  | 0.057 | **0.007** |
| **PFOA** | 0.016 | 0.652 |  | 0.021 | 0.639 |  | 0.040 | 0.494 |
| **PFNA** | 0.028 | 0.162 |  | 0.027 | 0.272 |  | 0.048 | 0.140 |
| **PFDA** | -0.005 | 0.790 |  | -0.003 | 0.879 |  | -0.008 | 0.772 |
| **PFUdA** | 0.009 | 0.505 |  | 0.020 | 0.226 |  | -0.011 | 0.596 |
| **PFHxS** | 0.000 | 0.999 |  | -0.010 | 0.706 |  | 0.036 | 0.291 |
| **PFOS** | 0.011 | 0.701 |  | 0.032 | 0.386 |  | 0.004 | 0.932 |
| **GenX** | 0.032 | 0.324 |  | 0.028 | 0.487 |  | 0.074 | 0.157 |
| **PFOSA** | 0.001 | 0.942 |  | 0.006 | 0.591 |  | -0.005 | 0.721 |
| **Total PFAS** | 0.021 | 0.541 |  | 0.026 | 0.546 |  | 0.058 | 0.302 |
| **NASEM PFAS^d^** | 0.023 | 0.546 |  | 0.045 | 0.340 |  | 0.031 | 0.624 |

a. CB: Cholesterol biomarker
b. β = change in mmol of cholesterol biomarker per ln-ng PFAS increase in serum.
c. Multivariable linear regression
